# Prevalence of early neonatal mortality and its predictors in sub-Saharan Africa: A Systematic review and Meta-Analysis

**DOI:** 10.1101/2024.08.06.24311554

**Authors:** Teebeny Zulu, Choolwe Jacobs, Godfrey Biemba, Patrick Musonda

## Abstract

**Background:** Although early neonatal mortality (ENM) has been reported to have a greater contribution to the overall neonatal mortality compared to late neonatal mortality, no meta-analysis has studied this phenomenon in isolation. The prevalence of ENM and its predictors in sub-Saharan Africa (SSA) remains unknown. Therefore, this meta-analysis is aimed at pooling the prevalence of ENM and its predictors in SSA.

**Methods:** Google Scholar, PubMed, Scopus, CINAHL, and Google were searched for studies conducted in SSA that reported the prevalence and predictors of ENM. The data were extracted using a Microsoft Excel spreadsheet and imported into R version 4.4.1 for further analysis. Publication bias, heterogeneity, sensitivity analysis, and subgroup analysis were performed. Prevalence and odds ratios were pooled using the random effects model if significant heterogeneity existed; otherwise, the fixed effects model was used.

**Results:** A total of 26 studies were included in this systematic review and meta-analysis. The overall pooled prevalence of ENM in SSA was 11% (95% CI: 7-15; *I^2^*=100%). Birth asphyxia (OR=3.85; 95% CI: 1.12-13.21; P = 0.0388; *I^2^* = 86.6%), home delivery (OR=2.46; 95% CI: 1.79-3.38; p<0.001; *I^2^* = 0.0%), prematurity (OR=4.69; 95% CI: 3.57-6.16; p<0.001; *I^2^* = 36.8%), male gender (OR= 1.37; 95% CI: 1.28-1.46; P < 0.001; *I^2^* = 30.7%), delivery through caesarean section (OR=1.74; 95% CI: 1.49-2.02; P < 0.001; *I^2^* = 31.5%) and low birth weight (OR=3.00; 95% CI: 1.01-8.91; P = 0.0482; *I^2^* = 94.4%) were associated with a significant increase in pooled odds of ENM in SSA.

**Conclusion:** The prevalence of ENM in SSA in significantly high and it contributes greatly to the overall neonatal mortality. Therefore, tailor-made interventions that target the reduction of birth asphyxia, prematurity, home delivery, and low birth weight should be implemented in order to reduce the burden of ENM in SSA.

## Introduction

Early neonatal mortality is defined as the death of a newborn that occurs within seven days of life [1]. Although neonatal mortality has decreased significantly over the past few decades, it still posesv a serious problem in most parts of the world [2]. Early neonatal mortality accounts for around 33% of all under-five deaths globally [3]. Of the 2.8 million babies that die throughout the neonatal period (0-28 days) globally each year, 73% do so in the first seven days of life [1, 4]. Ninety-nine percent of newborn fatalities and stillbirths take place in low- and middle-income countries like those in SSA [5]. Sub-Saharan Africa had the highest newborn mortality rate in 2019 with 27 deaths per 1,000 live births, followed by Central and Southern Asia with 24 deaths per 1,000 live births [6]. A child born in SSA or southern Asia had a tenfold higher probability of dying in the first month of life than a child born in a high-income country [6]. Sustainable Development Goal (SDG) 3 aims to reduce the neonatal mortality rate to 12 or less per 1000 live births by 2030 [7, 8]. Even though neonatal mortality has decreased significantly since 1990, further efforts are required to accelerate this progress and meet the SDG objective by 2030 [9].

Literature has shown that birth asphyxia [10–12], delivery through caesarean section [13–15], gestation less than 37 weeks [11, 13], giving birth from home [13, 16, 17], low birth weight [13, 15, 17, 18], male gender [16, 19], not attending antenatal care [11, 13], having no formal education [13, 16, 20] and prematurity [10, 21, 22] are factors associated with an increase in ENM.

The early neonatal period, the first seven days of life, is the most precarious for a baby’s survival and it contributes a higher percentage to the overall neonatal mortality [6]. Children face the highest risk of death in their first week of life [6], especially in low- and middle-income countries such as those of SSA. Despite this phenomenon being well documented in the literature, no meta-analysis has been performed to pool the evidence specifically, on the prevalence and predictors of ENM across individual studies conducted in SSA. An understanding of the burden and predictors of ENM is key in the development and sustainability of interventions aimed at reducing ENM in SSA. Therefore, this study provides a timely meta-analysis to understand the prevalence and predictors of ENM in SSA in order to bridge the existing gap in knowledge.

## Methods

### 2.1. Eligibility Criteria and Information Sources

This systematic review and meta-analysis included studies conducted in SSA to assess the prevalence of ENM and its predictors. The studies were evaluated using study area, study setting, title, abstract, and full texts before inclusion in this meta-analysis. This study is prepared based on preferred reporting items for systematic reviews and meta-analysis (PRISMA) statements [23]. In this review, published articles, surveys, and unpublished articles reported in English were explored and included accordingly. Studies that were conducted from inception to July 2024 were searched. Mendeley reference manager was used to manage retrieved articles.

### 2.2. Search Strategy and Selection of Studies

We conducted a comprehensive search to identify studies. Electronic databases, grey literature sources, and reference lists of articles were explored independently by three investigators Teebeny Zulu (TZ) Patrick Musonda (PM) and Godfrey Biemba (GB). Google Scholar, PubMed, Scopus, CINAHL, and Google were explored. Searching was conducted using key terms: (a) population (early neonates, perinatal, and newborns); (b) exposure (associated factors, risk factors, determinants, and predictors); (c) outcome (prevalence of early neonatal mortality, death, mortality, and birth outcomes); (d) study setting (hospitals, neonatal intensive care units, NICUs, facility-based, community-based surveys, and health centres); and (e) location (Sub-Saharan Africa, SSA, and Sub-Saharan countries). Boolean operators such as “OR” and “AND” were used during the search process. Screening and selection of studies was done using the Covidence screening tool by two authors (TZ and GB) and disagreements between the two authors was resolved by the third author (PM).

### 2.3. Data Extraction Process

A structured and pretested data extraction checklist in Microsoft Excel was used to extract information by two authors (TZ and GB). From included studies, we extracted the name of the first author, publication year, study country, sample sizes, study setting, study design, prevalence of ENM, adjusted odds ratio and their corresponding confidence intervals as the measure of association of the predictors.

### 2.4. Assessment of Study Quality

The Joanna Briggs Institute Prevalence Critical Appraisal Tool for use in the systematic review of prevalence studies was used for the critical appraisal of studies [24]. Moreover, the methodological and other qualities of each article were assessed based on a modified version of the Newcastle-Ottawa Scale adapted from [25].

### 2.5. Summary Measures

The primary outcome of this study was the prevalence of ENM (death before 7 days of life) in SSA. The second outcome was predictors of ENM that were computed from studies reporting predictors in the form of odds ratios. The pooled odds ratios were computed using the adjusted odds ratios with 95% confidence intervals (CI) that were reported in the original studies. The pooled odds ratios were presented with a 95% CI. Independent predictors were declared when p-value<0.05. The effect sizes were the prevalence of ENM and odds ratios predicting ENM.

### 2.6. Statistical Methods and Analysis

In the present meta-analysis, R software version 4.4.1 was used for computing the pooled estimates of both the odds ratios of ENM and its predictors. We used the *‘metaprop’* function from the Meta package for pooling the prevalence of ENM in SSA and the *‘metagen’* function for pooling log-odds ratios. The inverse method and the Der Simonian-Laird (DL) were used for pooling the prevalence and the heterogeneity parameter (Tau), respectively, in the random effects model. Subgroup analyses were performed using study setting (facility or community-based studies), region, and study design. Sensitivity analysis was performed using the leave-one-out method, which assesses the influence of each study on the pooled effect. Meta-analyses were presented using forest plots, summary tables, and texts.

### 2.7. Publication Bias and Heterogeneity

Publication bias was assessed by looking at the asymmetry of the funnel plot and/or the statistical significance of Egger’s regression test. Publication bias was declared when Egger’s regression test was significant (p < 0.05) [26]. Heterogeneities among studies were explored using forest plots, the I-squared test (*I^2^*), and Cochrane Q statistics [27]. The *I^2^* values of 25%, 50%, and 75% were interpreted as low, medium, and high heterogeneity, respectively [28]. In this study, the presence of heterogeneity was declared and justified when *I^2^* ≥ 50% with a p-value < 0.05. The sources of possible significant heterogeneities were explored through subgroup analyses and sensitivity analyses using the leave-one-out plot.

## 3. Results

### 3.1. Selection of Eligible Studies

In the initial search, 1676 studies were found. These studies were retrieved from electronic databases and other sources. Of these studies, 1326 were duplicate files, and 257 studies were removed after screening using abstracts. The full texts of 93 studies were reviewed. Finally, 26 studies [3, 10–22, 29–40] were included in the final analysis of this systematic review and meta-analysis (Figure 1).

**Figure 1:**
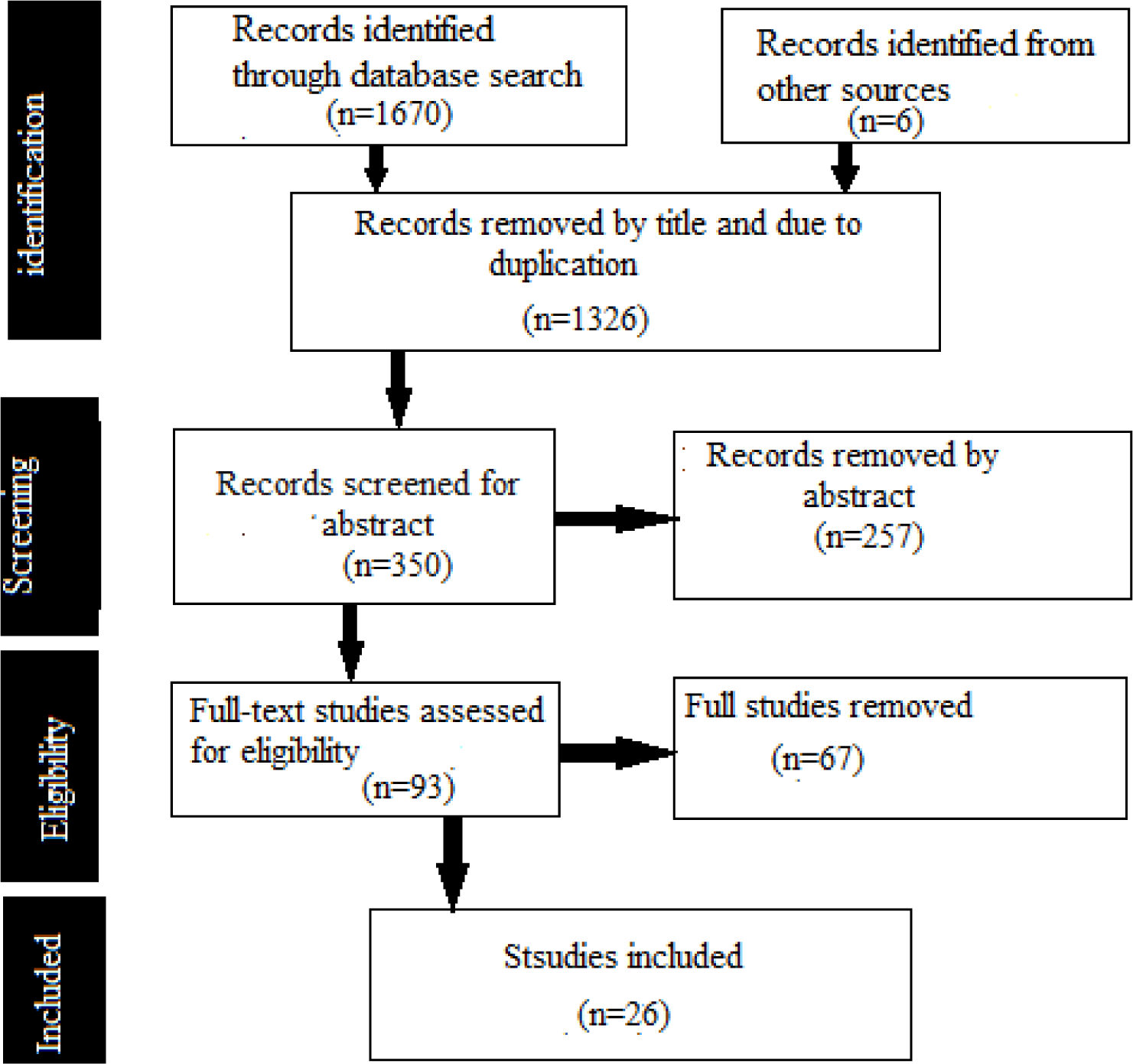
PRISMA flow diagram for showing screening and selection process of studies.

### 3.2. Characteristics of the Studies

Most articles in this systematic review and meta-analysis were conducted in Ethiopia (14). This meta-analysis included 18 studies from East Africa, 4 from West Africa, 2 from Central Africa, and 2 from selected countries in SSA. Sixteen studies were conducted in facilities, while 10 were conducted in communities. This meta-analysis consisted of 12 cross-sectional studies, 6 prospective cohort studies, 4 case-control studies, and 4 retrospective cohort studies. The proportion of ENM found in the studies varied from 1.5% to 33% as shown in Table 1.

**Table 1.**
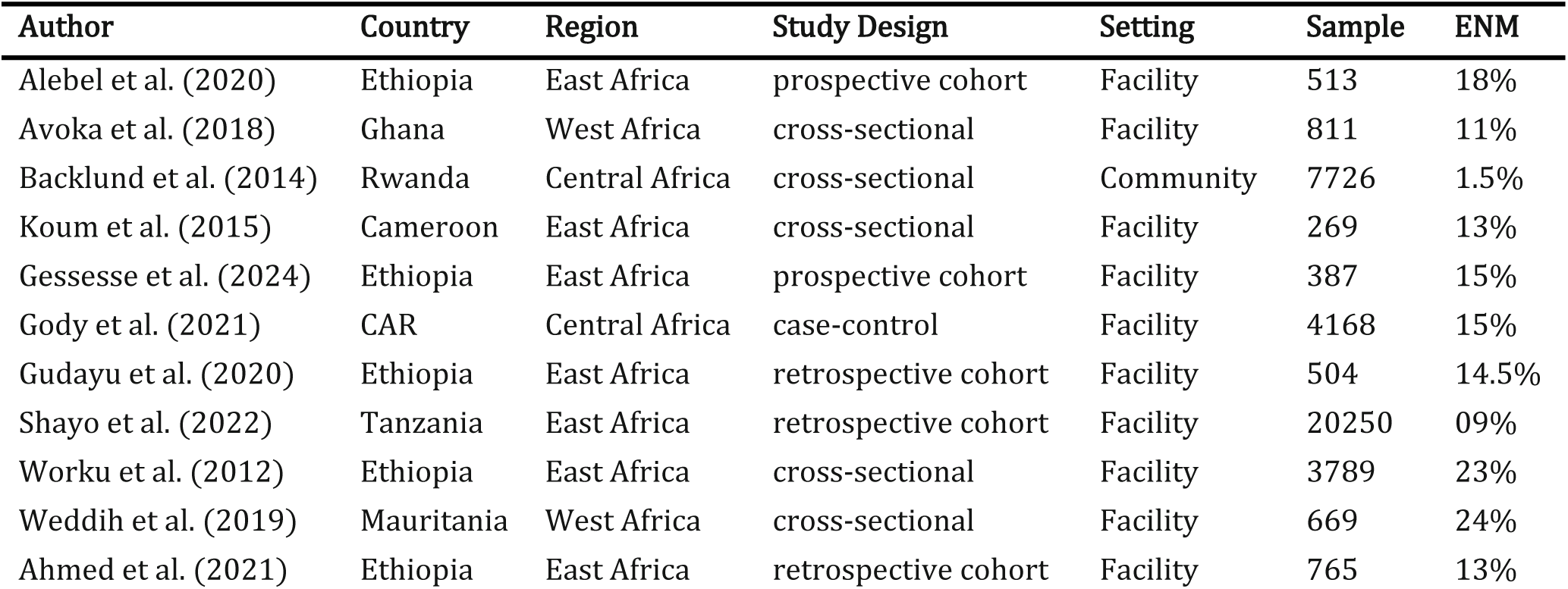
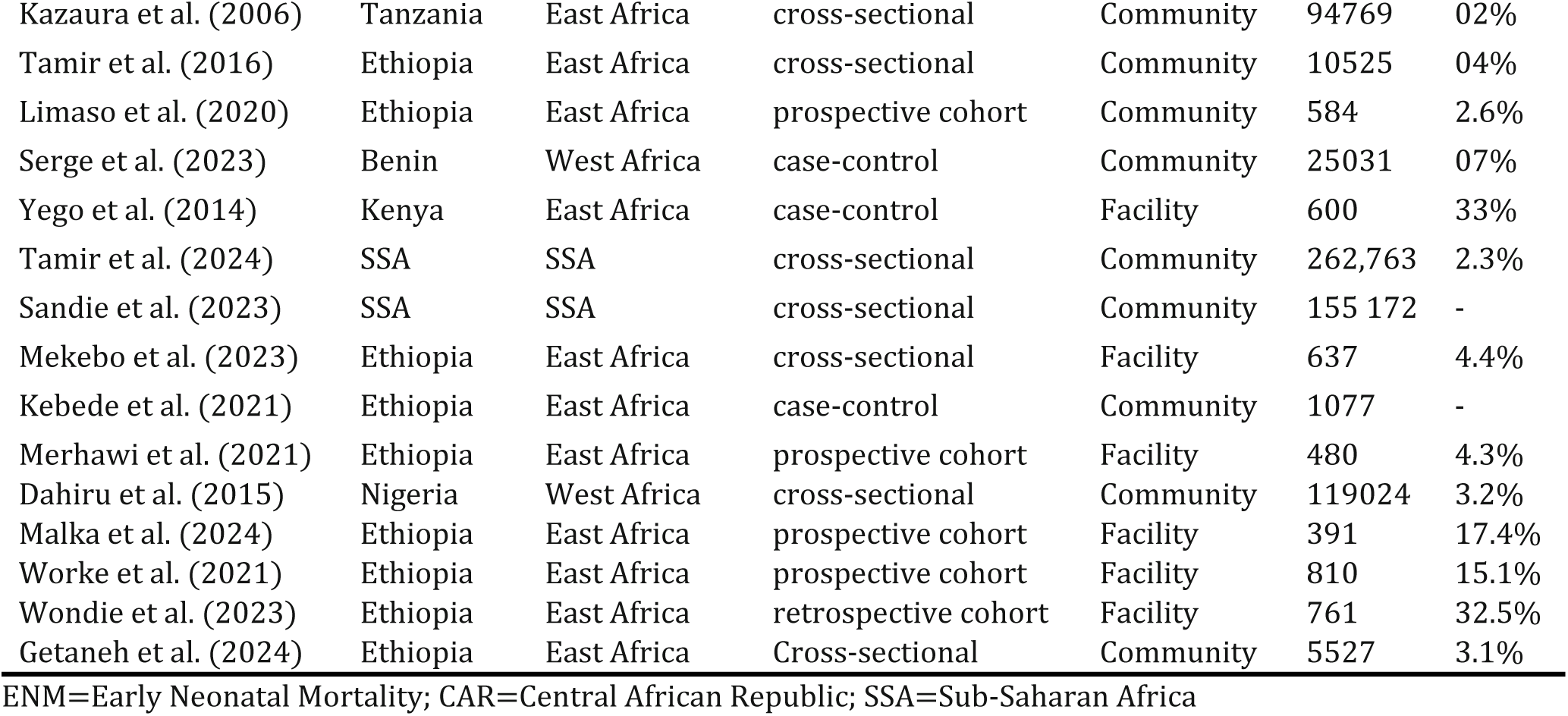
Characteristics of included studies for prevalence of ENM in SSA.

### 3.3. The Pooled prevalence of ENM in SSA

Twenty-three articles [3, 10-13, 16-21, 29-40] were included to estimate the pooled prevalence of ENM in SSA. The pooled prevalence of ENM in SSA was found to be 11%, or 110 deaths per 1000 live births (95% CI: 7%–15%; *I^2^*=100%) (Figure 2).

**Figure 2:**
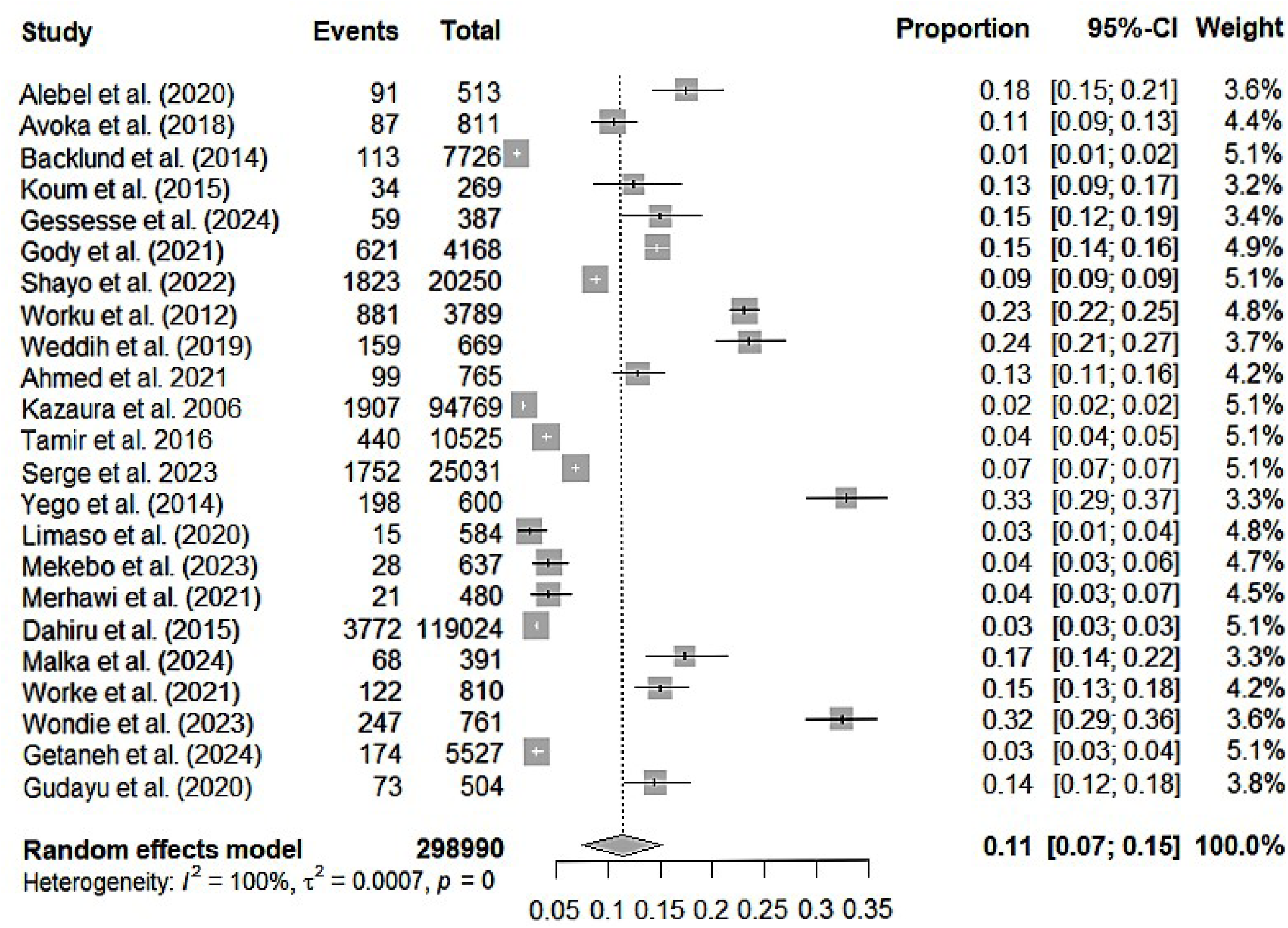
Forest plot for the pooled prevalence of ENM in SSA.

#### Subgroup analysis, publication bias and sensitivity analysis

Subgroup analysis was conducted on the basis of region, setting, and study design. Our regional subgroup analysis indicated that studies conducted in East Africa had the highest pooled prevalence of ENM at 13% (95% CI: 8%– 19%, *I^2^*=99%). (Figure 2). Figure 3 demonstrates that research done in facility settings showed a higher pooled prevalence of ENM than community-based studies, at 17% (95% CI: 14%–21%; *I^2^*=98%). Moreover, the greatest pooled prevalence 18% (95% CI: 10%-26%, *I^2^*=99%) came from case-control studies (Figure 4).

**Figure 3:**
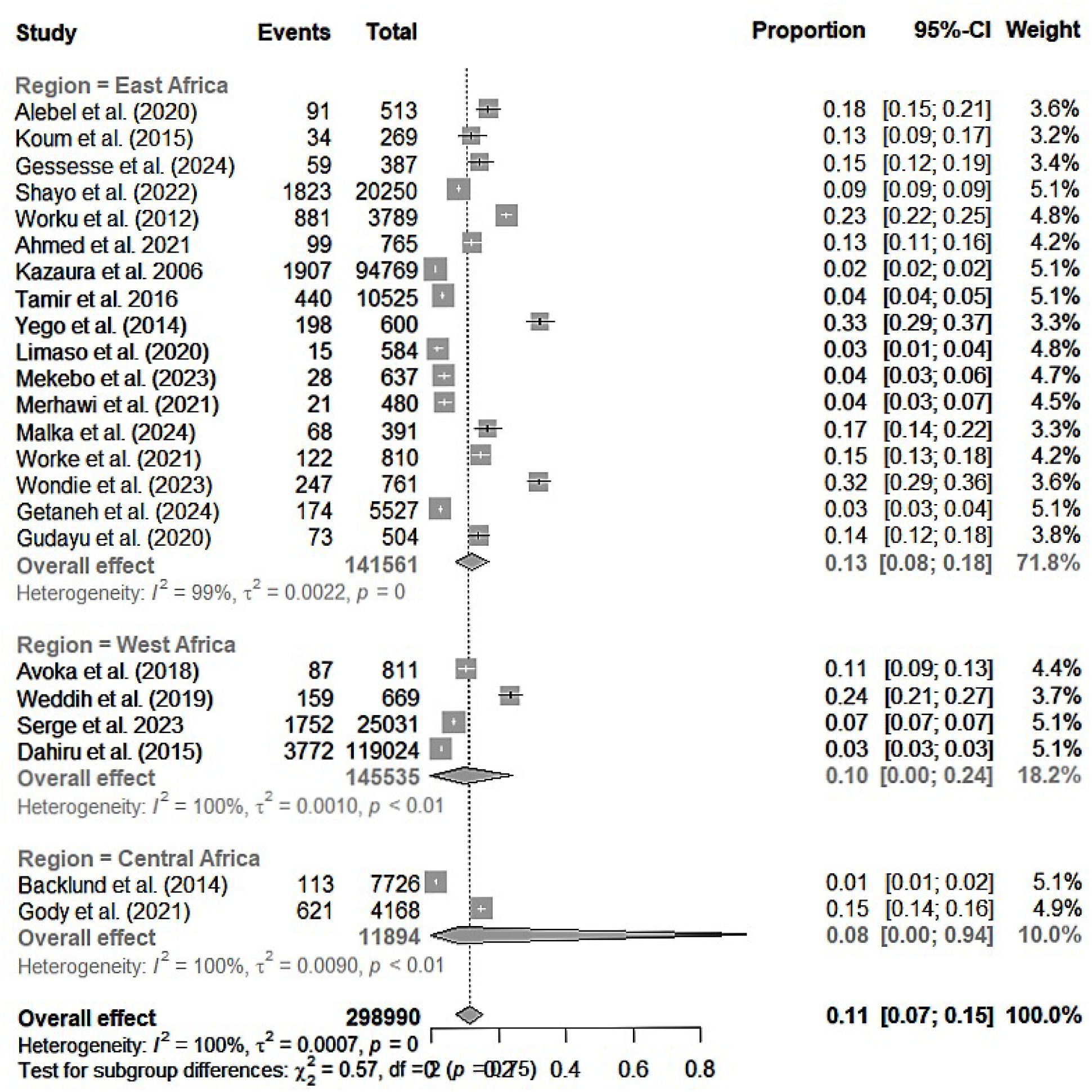
forest plot for subgroup analysis by study region.

**Figure 4:**
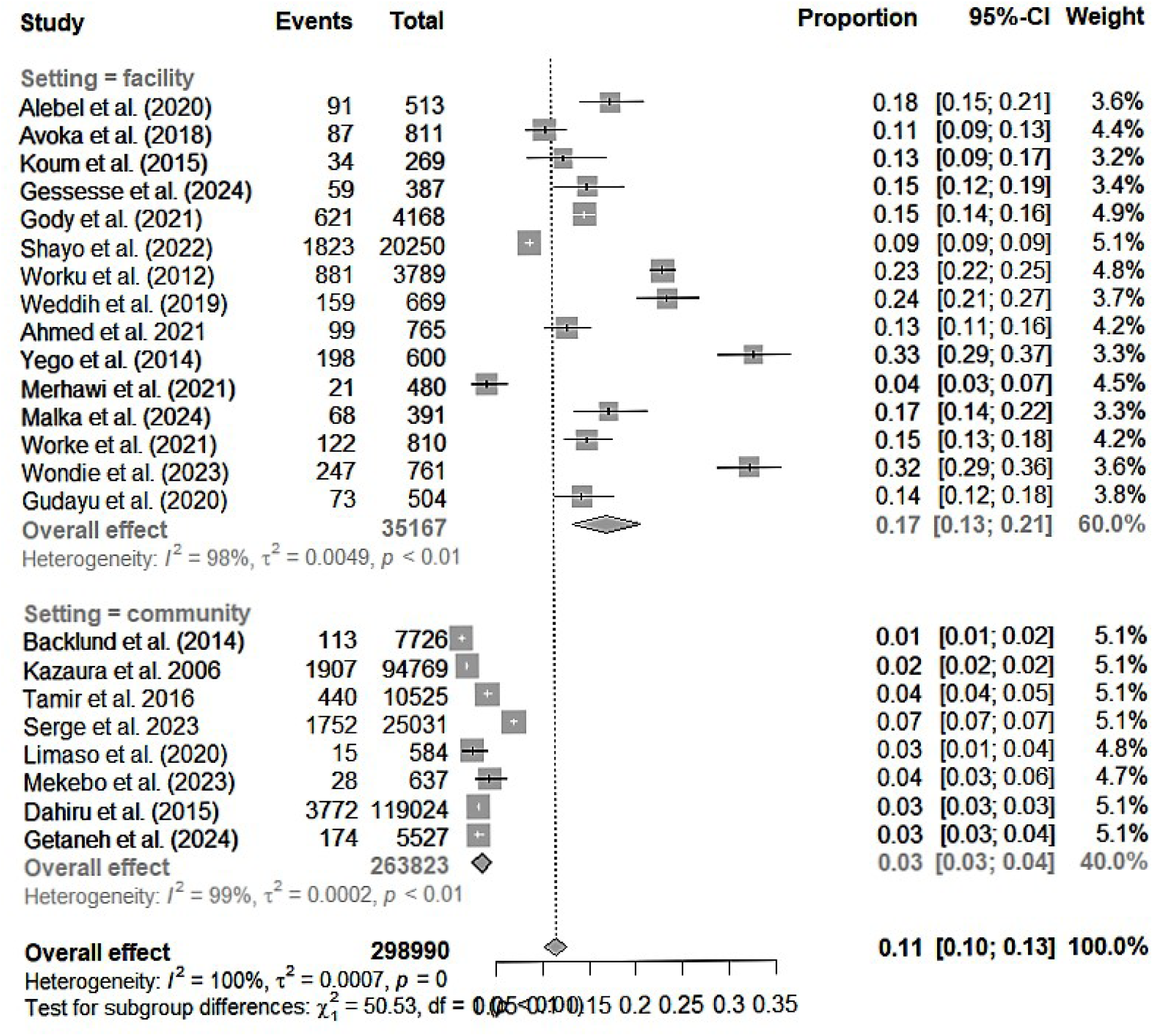
Forest plot for subgroup analysis by study setting.

An assessment of publication bias was done through a funnel plot and Egger’s regression test. Skewed distribution on a funnel plot suggested potential publication bias. Additionally, Egger’s regression test (p<0.001) objectively showed evidence of publication bias. Analysis utilizing the Der Simonian-Laird random-effects model revealed that no any individual study had an impact on the overall prevalence of ENM in SSA (Figure 7).

### 3.4 Factors associated with ENM in SSA

#### 3.4.1. Birth asphyxia

Five studies [10–13, 21] examined the association between birth asphyxia and ENM in SSA. The pooled odds of ENM for neonates who experienced birth asphyxia were 3.85 times the odds of those who did not experience it (95% CI: 1.12-13.21; *p*=0.0388) and heterogeneity (*I^2^* = 86.6%, *p* < 0.001).

**Figure 5:**
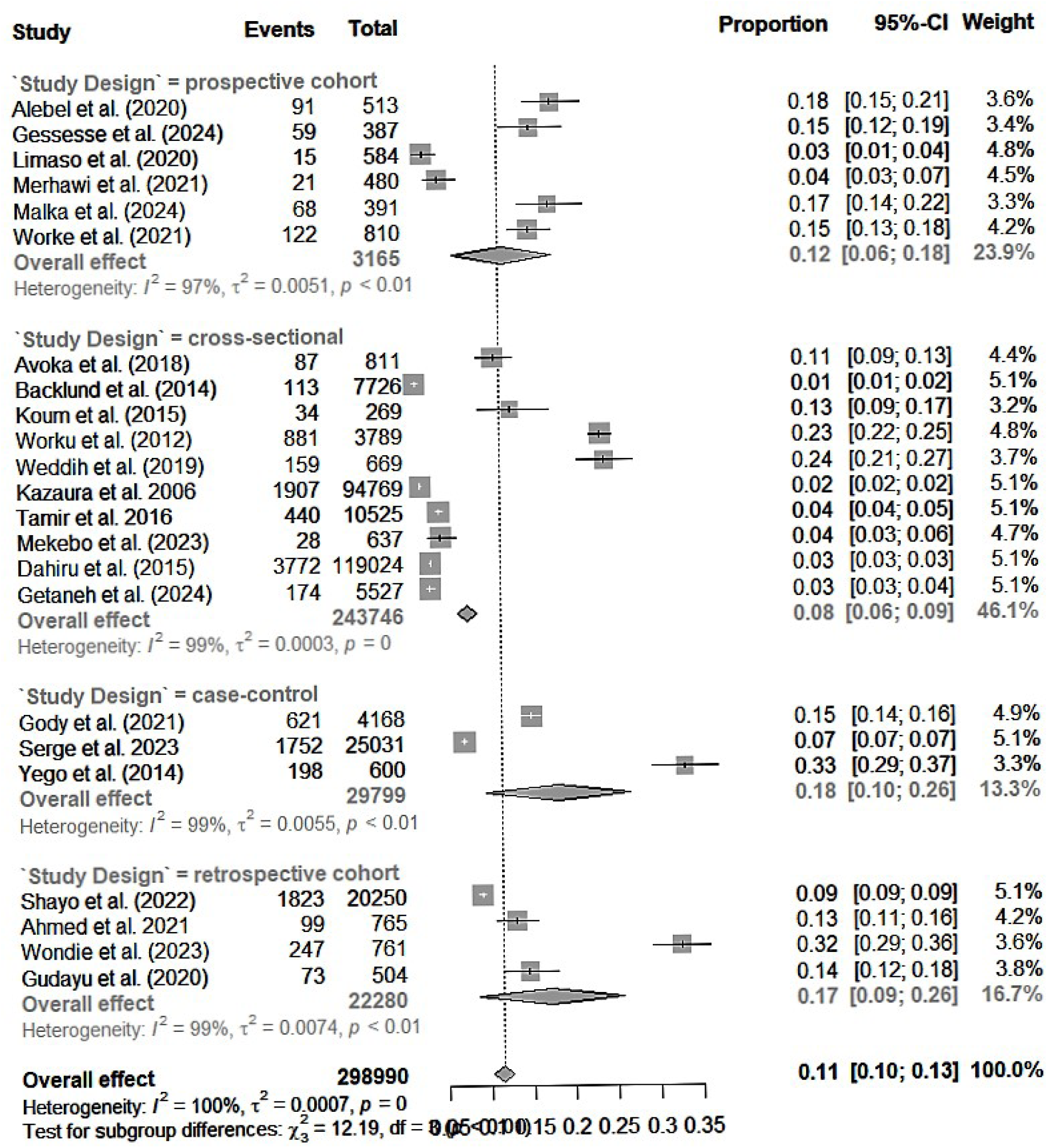
Forest plot for subgroup analysis by study design.

**Figure 6:**
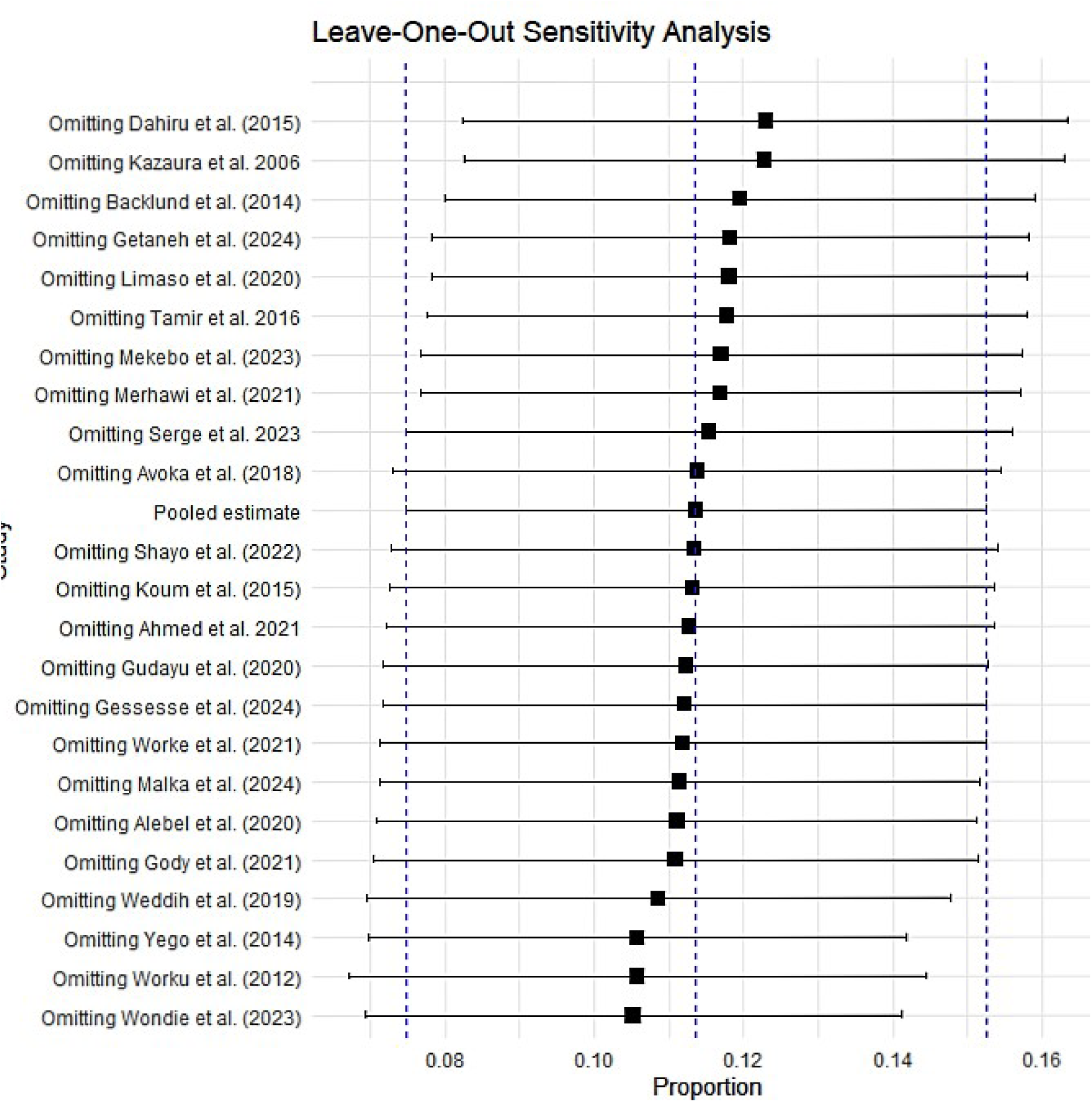
Sensitivity analysis stratified by effect size for studies included to estimate the pooled prevalence of ENM in SSA.

#### 3.4.2 Home delivery

Six studies [13, 15-17, 21, 31] examined the association between home delivery and ENM in SSA. The pooled odds of mortality among early neonates who were delivered at home were 2.46 times higher than their counterparts who were delivered at a health facility (95% CI: 1.79-3.38; p=0.0388). No significant heterogeneity was observed among these studies (*I*^2^ = 0.0%, *p* < 0.4305).

#### 3.4.3. Prematurity

We included 4 studies [10, 13, 21, 22] to assess the association between prematurity and ENM in SSA. The pooled odds of death for premature early neonates increased by a factor of 4.69 compared to their full-term counterparts (95% CI: 3.57-6.16; p<0.001) and heterogeneity (*I^2^* = 36.8%, *p* = 0.1915).

#### 3.4.4. Male gender

We included 8 studies [13, 14, 16, 19, 20, 31, 33, 36] to determine the association between male gender and ENM in SSA. As compared to female early neonates, the pooled odds of death for male early neonates increased by 37%, and this increase can be as low as 28% to as high as 46% in SSA with a 0.95 probability. No significant heterogeneity was observed among these studies (*I^2^* = 30.7%, *p* = 0.1831).

#### 3.4.5. Delivery through caesarean section

Eight studies [12–15, 17, 20, 22, 33] reported the association between delivery through a caesarean section and ENM in SSA. The pooled odds of ENM for neonates who were delivered through caesarean section were 74% higher than those who were delivered through the vagina (95% CI: 1.49–2.02; *p* < 0.001) and heterogeneity (*I^2^* = 31.5%, *p* = 0.1768).

#### 3.4.6. Low birth weight (>2.5kg)

The odds ratio of the association between low birth weight and ENM was pooled from 13 studies [11-15, 17-21, 30, 31, 36]. Low birth weight was found to increase ENM by a factor of 3 compared to normal birth weight (95% CI: 1.01-8.91; p<0.0482) and heterogeneity (*I^2^* = 94.4%, p < 0.001).

On the other hand, having no education compared to having some education (OR = 1.44; 95% CI: 0.72-2.89, *P* = 0.2454, *I^2^* = 94.4%), not attending antenatal care compared to attending (OR = 3.56; 95% CI: 0.64-19.84, *P* = 0.0864, *I^2^* = 93.1%), having respiratory distress compared to not having (OR = 4.73; 95% CI: 0.01-2170.45, *P* = 0.3893, *I^2^* = 83.9%), and gestation less than 37 weeks (OR = 12.49; 95% CI: 0.85-182.94, *P* = 0.0560, *I^2^* = 91.1%) were associated with an increase in the pooled odds of ENM in SSA, though the association was not statistically significant.

## 4. Discussion

This meta-analysis was aimed at estimating the pooled prevalence of ENM and its associated predictors in SSA. The study found a relatively high prevalence of ENM in SSA. Birth asphyxia, home delivery, prematurity, male gender, delivery through a caesarean section, and low birth weight were factors associated with a significant increase in neonatal mortality.

### 4.1 prevalence of ENM in SSA

Our meta-analysis is one of its kind, pooling evidence from studies across SSA to explore the prevalence and predictors of ENM in SSA. The pooled prevalence of ENM in SSA was estimated from 23 studies, and it was found to be 11% (110 deaths per 1000 live births). The high prevalence observed is not surprising as a higher percentage of newborn deaths occur in the first week of life especially in limited resource settings like SSA [1, 5]. The high prevalence can also be explained by the large number of facility-based studies included in this meta-analysis which overestimates ENM. Subgroup analysis revealed that East Africa reported the highest prevalence of ENM in SSA (13%). We also found that the pooled prevalence of ENM was higher in facility-based studies (17%) as compared to community-based studies (3%), mostly those performed on national demographic and health surveys. These figures show that the prevalence of ENM is still high, and there is a need to scale up efforts aimed at reducing the burden of ENM in SSA.

### 4.2 Predictors of ENM in SSA

Our meta-analysis combined odds ratios for 10 predictors of ENM in sub-Saharan Africa. A predictor had to be included in an adjusted model in a minimum of three studies to be included in the meta-analysis. We observed that birth asphyxia, delivering at home, being born prematurely, being male, having a caesarean section, and having a low birth weight were significantly linked to an increase in ENM. However, not having received a formal education, not attending antenatal care, experiencing respiratory distress, and having a gestation period of less than 37 weeks were found to be factors that increased the risk of ENM. However, the combined odds ratios for these factors did not show statistical significance in this research.

Our research revealed that newborns aged 0-7 days who suffered from birth asphyxia had a higher chance of mortality compared to those who didn’t undergo such a condition. This result aligns with the results of previous studies [10–13, 21]. Birth asphyxia occurs when a newborn’s brain and other organs are deprived of sufficient oxygen and nutrients before, during, or immediately following delivery. Birth asphyxia is one of the leading causes of mortality for newborns; the effect of birth asphyxia is not limited to death but also leads to physical, mental, and social incapability in newborns due to severe hypoxic-ischemic organ damage [41]. Hence, to reduce overall newborn mortality and its long-term consequences, the quality of medical care before birth, at birth, and after birth is essential.

**Figure 7:**
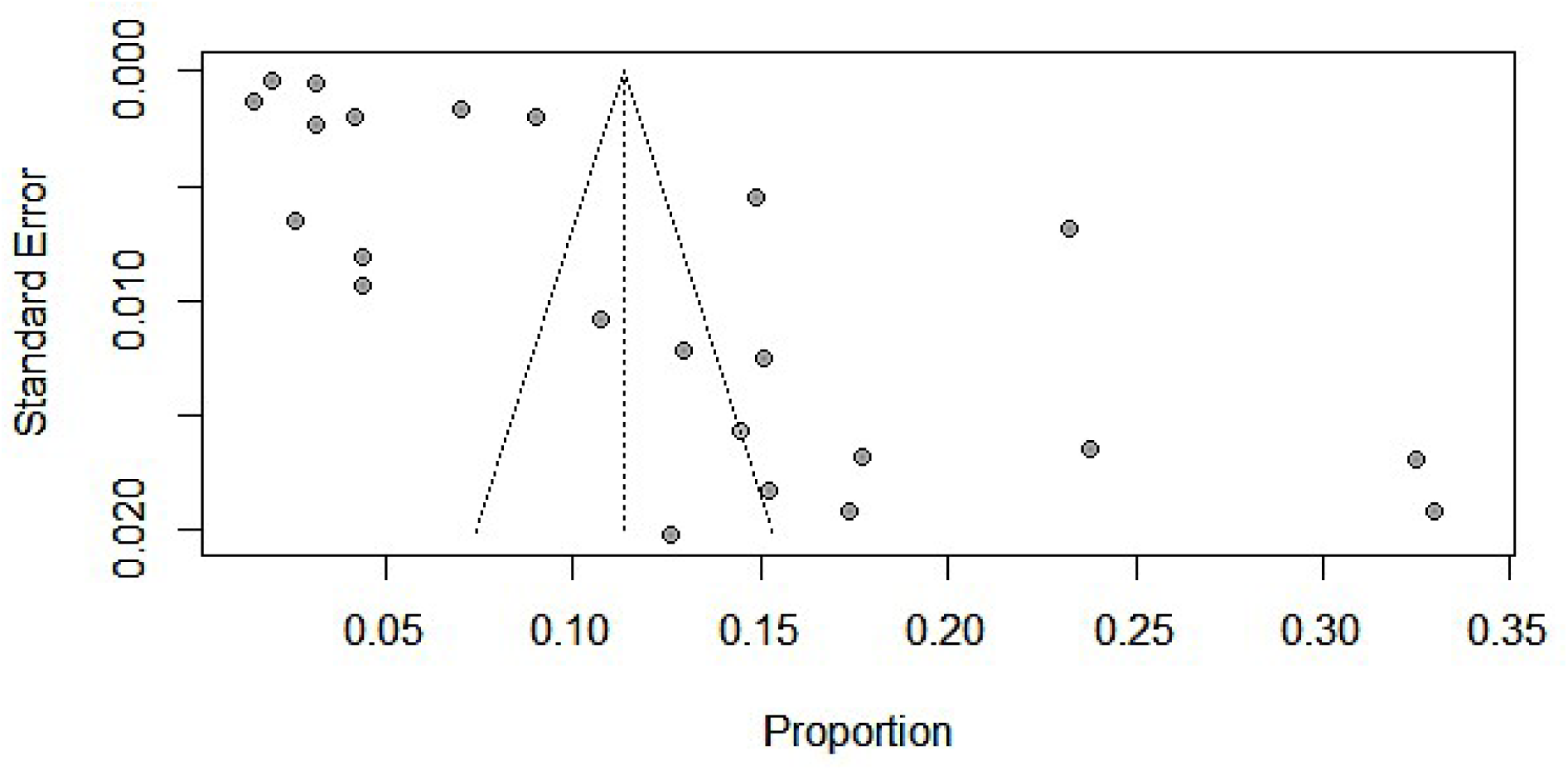
Funnel plot showing publication bias among studies included to estimate a pooled prevalence of ENM in SSA.

Findings have shown that newborn babies born at home had a higher probability of dying within the first week of life than those born in a medical center. This discovery aligns with findings from other research studies [13, 16, 17, 21, 42]. Babies born at home with a non-professional attendant may not receive all necessary newborn and postpartum care services. In cases of delivery complications, it is best to opt for institutional deliveries as both mothers and babies receive professional care and have access to life-saving tools at healthcare facilities, resulting in mutual benefits. Further, having a skilled team at a healthcare facility could provide an additional benefit in minimizing possible delays and improving the early treatment of pregnancy-related issues and birth difficulties [43]. This discovery shows a necessity to enhance the provision of health facility delivery in SSA in order to raise the likelihood of newborn survival. Male babies were more likely to die within the first week of life after birth compared to female babies. This result is consistent with the previous studies [16, 19, 33]. The possible reason could be that males are biologically weaker than females. Therefore, specialized care should be given to newborns especially, male ones.

**Table 2:** predictors of ENM in SSA.

| Predictors | Included studies | OR (95% CI) | POR (95% CI), p-value | Heterogeneity |
| --- | --- | --- | --- | --- |
| ***Birth asphyxia*** | Ahmed et al. 2021<br>Koum et al. 2015<br>Gody et al. 2021<br>Worku et al. 2012<br>Yego et al. 2014 | 2.20 (0.94, 5.13)<br>17.80 (2.01, 157.42)<br>12.72 (6.54, 24.73)<br>1.82 (1.32, 2.51)<br>2.40 (1.60, 3.60) | 3.85 (1.12, 13.21)<br>**P = 0.0388** | $I^2 = 86.6\%$,<br>$p < 0.001^{**}$ |
| ***Home delivery*** | Ahmed et al. 2021<br>Backlund et al. 2014<br>Gody et al. 2021<br>Mekebo et al. 2023<br>Tamir et al. 2024<br>Tamir et al. 2016 | 1.00 (0.20, 4.95)<br>3.50 (1.89, 6.49)<br>1.43 (0.64, 3.20)<br>2.29 (0.97, 5.39)<br>3.59 (1.41, 9.14)<br>2.40 (1.32, 4.36) | 2.46 (1.79, 3.38)<br>**P < 0.001** | $I^2 = 0.0\%$,<br>$p = 0.4305^*$ |
| ***Prematurity*** | Ahmed et al. 2021<br>Koum et al. 2015<br>Gody et al. 2021<br>Kebede et al. 2021 | 3.60 (1.66, 7.81)<br>21.30 (1.45, 313.96)<br>6.11 (4.09, 9.12)<br>3.62 (2.36, 5.55) | 4.69 (3.57, 6.16)<br>**p < 0.001** | $I^2 = 36.8\%$,<br>$p = 0.1915^*$ |
| ***Male gender*** | Backlund et al. 2014<br>Gody et al. 2021<br>Kazaura et al. 2006<br>Mekebo et al. 2023<br>Sandie et al. 2023<br>Shayo et al. 2022<br>Serge et al. 2023 | 1.43 (0.86, 2.38)<br>1.27 (0.88, 1.84)<br>1.40 (1.30, 1.50)<br>1.63 (0.79, 3.36)<br>1.23 (0.94, 1.62)<br>1.50 (1.14, 1.98)<br>1.04 (0.68, 1.59) | 1.38 (1.30, 1.48)<br>**P < 0.001** | $I^2 = 0.0\%$,<br>$p = 0.7703^*$ |
| ***Delivery through C-section*** | Gody et al. 2021<br>Kebede et al. 2021<br>Sandie et al. 2023<br>Shayo et al. 2022<br>Tamir et al. 2024<br>Tamir et al. 2016<br>Serge et al. 2023<br>Yego et al. 2014 | 1.69 (1.18, 2.42)<br>1.16 (0.78, 1.72)<br>2.37 (1.64, 3.42)<br>2.10 (1.48, 2.98)<br>1.81 (1.31, 2.51)<br>1.60 (0.69, 3.73)<br>1.13 (0.64, 2.00)<br>1.50 (0.29, 7.70) | 1.74 (1.49, 2.02)<br>**P < 0.001** | $I^2 = 31.5\%$,<br>$p = 0.1768^*$ |
| ***Low birth weight*** | Ahmed et al. 2021<br>Avoka et al. 2018<br>Backlund et al. 2014<br>Gody et al. 2021<br>Kazaura et al. 2006<br>Sandie et al. 2023<br>Tamir et al. 2024<br>Tamir et al. 2016<br>Serge et al. 2023<br>Worku et al. 2012<br>Yego et al. 2014<br>Weddih et al. 2019 | 2.50 (0.30, 21.16)<br>2.0700 (1.09, 3.92)<br>3.45 (1.88, 6.34)<br>22.59 (15.93, 32.04)<br>18.40 (15.81, 21.41)<br>2.35 (0.50, 10.94)<br>3.27 (2.16, 4.95)<br>3.30 (1.36, 7.99)<br>0.84 (0.30, 2.35)<br>9.64 (3.32, 27.98)<br>2.40 (0.88, 6.54)<br>3.91 (1.69, 9.05) | 4.29 (2.30, 8.02)<br>**P = 0.0003** | $I^2 = 94.0\%$,<br>$p < 0.001^{**}$ |

**Table 2 continued**
| Predictors | Included studies | OR (95% CI) | POR (95% CI), p-value | Heterogeneity |
| --- | --- | --- | --- | --- |
| *No education* | Gody et al. 2021<br>Kazaura et al. 2006<br>Mekebo et al. 2023<br>Sandie et al. 2023<br>Tamir et al. 2024<br>Tamir et al. 2016<br>Serge et al. 2023 | 5.65 (4.08, 7.82)<br>1.29 (0.66, 2.54)<br>2.13 (1.14, 3.99)<br>1.14 (0.88, 1.47)<br>0.83 (0.69, 1.00)<br>0.50 (0.17, 1.46)<br>1.30 (1.08, 1.56) | 1.44 (0.72, 2.89)<br>**P = 0.2454** | $I^2 = 94.4\%$<br>$p < 0.001^{**}$ |
| *No antenatal care visits* | Gody et al. 2021<br>Worku et al. 2012<br>Yego et al. 2014 | 5.54 (3.95, 7.78)<br>1.70 (1.28, 2.26)<br>5.40 (1.99, 14.64) | 3.56 (0.64, 19.84)<br>$P = 0.0864$ | $I^2 = 93.1\%$,<br>$p < 0.001^{**}$ |
| *Respiratory distress* | Koum et al. 2015<br>Gody et al. 2021<br>Yego et al. 2014 | 130.37 (9.28, 1831.92)<br>1.22 (0.88, 1.69)<br>1.60 (1.08, 2.36) | 4.73 (0.01, 2170.45) $P = 0.3893$ | $I^2 = 83.9\%$,<br>$p = 0.002^{**}$ |
| *Gestation less than 37 weeks* | Gody et al. 2021<br>Worku et al. 2012<br>Yego et al. 2014 | 29.36 (20.13, 42.83)<br>3.60 (1.64, 7.90)<br>16.60 (8.19, 33.65) | 12.49 (0.85, 182.94)<br>$P = 0.0560$ | $I^2 = 91.1\%$,<br>$p < 0.001^{**}$ |

OR=odds ratio; POR=pooled odds ratio CI=confidence interval; \*=fixed effects model; \*\*=random effects model

Consistent with previous research [21, 44], premature infants had a higher likelihood of mortality in the first week of life in comparison to full-term infants. Preterm babies face risks such as organ failure, neurodevelopmental and learning disabilities, vision problems, and long-term cardiovascular and non-communicable diseases [45]. Hence, it is necessary to introduce measures that can decrease early mortality rates related to prematurity. For instance, according to a systematic review of low and middle-income countries [46], giving pregnant mothers multiple micronutrient supplements and improving the quality of antenatal care can decrease prematurity and stillbirth. In pre-term neonates, feeding support, probiotics, and thermal regulation were reported to improve survival rates in premature newborns.

Although a cesarean section (CS) is performed to save the life of the newborn, the possibility of ENM was higher among babies delivered by CS compared to babies delivered vaginally. This finding was coherent with previous studies [15, 14, 47, 48]. On this basis, previous scientific literature also recommends avoiding CS as an intrapartum intervention when there is no clear medical indication that it will improve the outcome for the mother or the baby [44, 47]. However, it could be plausible that the higher likelihood of ENM among babies born via CS was since the majority of CS deliveries occur as a last option for delivery when there are pregnancy complications [15]. Another possible explanation for this finding in SSA could be due to poor labour care, poor surgical care at the time of cesarean section, and poor neonatal care post-CS that may be observed in most SSA countries’ health facilities [13].

Regarding birth weight, the likelihood of ENM was higher among low birth-weight (LBW) babies compared to babies of normal birth weight. This is consistent with previous studies [15, 49, 50]. One of the common reasons for the higher risk of ENM among LBW babies is that most of the time, LBW babies are preterm births and/or small for gestational age [49]. These findings suggest the need to improve mother care during pregnancy, childbirth, and postnatal periods, particularly for LBW babies. The World Health Organisation has also developed clinical guidelines to increase baby survival and advocates the need for careful essential newborn care for LBW babies [51]. Kangaroo mother care can be an option for LBW neonates which involves skin-to-skin contact between a mother and her newborn [52].

## Study strengths and limitations

The main strength of this study is that reputable databases were explored to find all possible articles. This study is the first of its type to determine the prevalence of ENM in SSA and unfold the possible predictors of ENM using odds ratios. In addition, the subgroup analysis performed in this study which revealed high prevalence among facility-based studies indicates the quality of newborn care service delivery across facilities in SSA. This finding will have a paramount importance for program planners and policymakers in SSA, where the burden of ENM is considerably high. Despite these strengths, the presence of scant studies in countries other than Ethiopia could obscure some other predictors of ENM in SSA. The lack of reports on the prevalence of ENM in community-based studies could also be associated with underestimation of the prevalence of ENM in SSA. Studies in countries other than Ethiopia and community-based studies could unfold all possible causes of ENM in the future.

## Conclusion

We determined the prevalence of ENM and its predictors in SSA. The pooled prevalence of ENM was significantly high in SSA. Facility-based studies contributed greatly to the high prevalence of ENM in SSA. Birth asphyxia, home delivery, prematurity, male gender, delivery through cesarean section and low birth weight were factors associated with a significant increase in the likelihood of ENM in SSA. Therefore, there is a need to design strategies to address all these predictors of ENM to reduce the high burden of ENM in SSA.

## Data Availability

citations to the studies used in this systematic review and meta-analysis are provided in this study and can be freely accessed.

## References

1. Lehtonen L., et al. Early neonatal death: a challenge worldwide in Seminars in Fetal and Neonatal Medicine. 2017. Elsevier. 10.1016/j.siny.2017.02.006 PMID: 28238633.

2. Kibria G.M.A., et al., Determinants of early neonatal mortality in Afghanistan: an analysis of the Demographic and Health Survey 2015. Globalization and health, 2018. 14(1): p. 1–12

3. Dahiru T. Determinants of Early Neonatal Mortality in Nigeria: Results from 2013 Nigeria DHS. J Pediatric Neonatal Care. 2015; 2 (5):1–8.

4. Lawn J.E., et al., Every Newborn: progress, priorities, and potential beyond survival. The lancet, 2014. 384(9938): p. 189–205. 10.1016/S0140-6736(14)60496-7 PMID: 24853593

5. Arsenault C., et al., Equity in antenatal care quality: an analysis of 91 national household surveys. The Lancet Global Health, 2018. 6(11): p. e1186–e1195. 10.1016/S2214-109X(18)30389-9 PMID: 30322649

6. Newborns, W., Improving survival and well-being. World Health Organization. https://www.who.int/news-room/fact-sheets/detail/newborns-reducing-mortality, 2020

7. Transforming our world Cf, O. The 2030 Agenda for Sustainable Development. United Nations: New York, NY, USA, 2015.

8. Sudfeld C.R. and Fawzi W.W., Importance of innovations in neonatal and adolescent health in reaching the sustainable development goals by 2030. JAMA pediatrics, 2017. 171(6): p. 521–522. 10.1001/jamapediatrics.2017.0261 PMID: 28384685

9. Hug L., et al., National, regional, and global levels and trends in neonatal mortality between 1990 and 2017, with scenario-based projections to 2030: a systematic analysis. The Lancet Global Health, 2019. 7(6): p. e710–e720. 10.1016/S2214-109X(19)30163-9 PMID: 31097275.

10. Koum DCK, Essomba NE, Moby H, Halle MPE, Ngaba GP, Nguedjam MM, et al. Factors associated with early neonatal morbidity and mortyality in an urban district hospital in Douala, Cameroon. Int J latest Res Sci Technol. 2016; 5(3):43–9.

11. Worku B, Kassie A, Mekasha A, Tilahun B, Worku A. Predictors of early neonatal mortality at a neonatal intensive care unit of a specialized referral teaching hospital in Ethiopia. Ethiop J Heal Dev. 2012; 26(3):200–7.

12. Yego F, D’Este C, Byles J, Nyongesa P, Williams JS. A case-control study of risk factors for fetal and early neonatal deaths in a tertiary hospital in Kenya. BMC Pregnancy Childbirth. 2014; 14(1):1–9.

13. Gody JC, Engoba M, Mejiozem BOB, Danebera LV, Kakouguere EP, Bangue MCN, et al. Risk Factors of Early Neonatal Deaths in Pediatric Teaching Hospital in Bangui, Central African Republic. Open J Pediatr. 2021; 11(04):840–53.

14. Sandie AB, Mutua MK, Sidze E, Nyakangi V, Sylla EHM, Wanjoya A, et al. Epidemiology of emergency and elective caesarean section and its association with early neonatal mortality in sub-Saharan African countries. BMJ Open. 2023; 13(10).

15. Tamir TT, Mohammed Y, Kassie AT, Zegeye AF. Early neonatal mortality and determinants in sub-Saharan Africa: Findings from recent demographic and health survey data. PLoS One [Internet]. 2024; 19(6):e0304065. Available from: 10.1371/journal.pone.0304065

16. Mekebo GG, Aga G, Gondol KB, Regesa BH, Woldeyohannes B, Wolde TS, et al. Why Babies die in the first 7 days after birth in Somalia Region of Ethiopia? Ann Med Surg. 2023; 85(5):1821–5.

17. Tamir TT, Asmamaw DB, Negash WD, Belachew TB, Fentie EA, Kidie AA, et al. Prevalence and determinants of early neonatal mortality in Ethiopia: Findings from the Ethiopian Demographic and Health Survey 2016. BMJ Paediatr Open. 2023; 7(1):1–8.

18. Weddih A, Ahmed MLCB, Sidatt M, Abdelghader N, Abdelghader F, Ahmed A, et al. Prevalence and factors associated with neonatal mortality among neonates hospitalized at the national hospital Nouakchott, Mauritania. Pan Afr Med J. 2019; 34:1–7.

19. Kazaura. R, Kidanto H LSN. M. Level,trends and risk for early neonatal mortality at Muhimbili National Hospital,Tanzania,199-2005. East African J Public Heal. 2006; 3(10):10–3.

20. Serge T, Georgia DB, Badirou A. Epidemiological Aspects and Factors Associated with Early Neonatal Death From 2018 to 2020 in the Maternity of the Savè-Ouessè Health Zone, Benin, West Africa. J Matern Child Heal. 2023; 8(1):91–104.

21. Ahmed AT, Farah AE, Ali HN, Ibrahim MO. Determinants of early neonatal mortality (hospital based retrospective cohort study in Somali region of Ethiopia). Sci Rep. 2023; 13(1):1–13.

22. Kebede E, Kekulawala M. Risk factors for stillbirth and early neonatal death: a case-control study in tertiary hospitals in Addis Ababa, Ethiopia. BMC Pregnancy Childbirth. 2021; 21(1):1–11.

23. Page MJ, McKenzie JE, Bossuyt PM, Boutron I, Hofmann TC, Mulrow CD, et al. The PRISMA 2020 Statement: an updated guideline for reporting systematic reviews. Bmj. 2021; 372.

24. Munn Z., S. Moola, K. Lisy, D. Riitano, and C. Tufanaru, “Methodological guidance for systematic reviews of observational epidemiological studies reporting prevalence and cumulative incidence data,” International Journal of Evidence-Based Healthcare, vol. 13, no. 3, pp. 147–153, 2015.

25. Modesti P. A., G. Reboldi, F. P. Cappuccio et al., “Panethnic differences in blood pressure in Europe: a systematic review and meta-analysis,” PLoS One, vol. 11, no. 1, article e0147601, 2016.

26. Sterne and M. Egger J. A. C., “Funnel plots for detecting bias in meta-analysis: guidelines on choice of axis,” Journal of Clinical Epidemiology, vol. 54, no. 10, pp. 1046–1055, 2001.

27. Rücker G, Schwarzer G, Carpenter J. R, and Schumacher M, “Undue reliance on I 2 in assessing heterogeneity may mislead,” BMC Medical Research Methodology, vol. 8, no. 1, 2008.

28. Higgins J. P. T, and Thompson S. G., “Quantifying heterogeneity in a meta-analysis,” Statistics in Medicine, vol. 21, no. 11, pp. 1539–1558, 2002.

29. Alebel A, Wagnew F, Petrucka P, Tesema C, Moges NA, Ketema DB, et al. Neonatal mortality in the neonatal intensive care unit of Debre Markos referral hospital, Northwest Ethiopia: A prospective cohort study. BMC Pediatr. 2020; 20(1):1–11.

30. Avoka JA, Adanu RM, Wombeogo M, Seidu I, Dun-Dery EJ. Maternal and neonatal characteristics that influence very early neonatal mortality in the Eastern Regional Hospital of Ghana, Koforidua: A retrospective review. BMC Res Notes [Internet]. 2018; 11(1):1–5. Available from: 10.1186/s13104-018-3196-x

31. Backlund A. Maternal health care in Rwanda and its association to early neonatal mortality. 2015;

32. Gessesse AD, Belete MB, Tadesse F. Dar City public hospitals. 2024; (June):1–16. Available from: 10.3389/fped.2024.1335858

33. Shayo A, Mlay P, Ahn E, Kidanto H, Espiritu M, Perlman J. Early neonatal mortality is modulated by gestational age, birthweight and fetal heart rate abnormalities in the low resource setting in Tanzania – a five year review 2015–2019. BMC Pediatr [Internet]. 2022; 22(1):1–11. Available from: 10.1186/s12887-022-03385-0

34. Wondie WT, Zeleke KA, Wubneh CA. Incidence and predictors of mortality among low birth weight neonates in the first week of life admitted to the neonatal intensive care unit in Northwestern Ethiopia comprehensive specialized hospitals, 2022. Multi-center institution-based retrospective f. BMC Pediatr. 2023; 23(1):1–13.

35. Worke MD, Mekonnen AT, Limenh SK. Incidence and determinants of neonatal mortality in the first three days of delivery in northwestern Ethiopia: a prospective cohort study. BMC Pregnancy Childbirth [Internet]. 2021; 21(1):1–11. Available from: 10.1186/s12884-021-04122-8.

36. Gudayu T. W, Zeleke E. G, and Lakew A. M, “Time to death and its predictors among neonates admitted in the intensive care unit of the University of Gondar Comprehensive Specialized Hospital, Northwest Ethiopia,” Research and Reports in Neonatology, vol. Volume 10, no. 1, pp. 1–10, 2020.

37. Merhawi B, G w H. Early Neonatal Death in Northern Ethiopia and its Predictors. Res Artic Divers Equal Heal Care. 2021; 18(11):488–92.

38. Malka ES, Solomon T, Kassa DH, Erega BB, Tufa DG. Time to death and predictors of mortality among early neonates admitted to neonatal intensive care unit of Addis Ababa public Hospitals, Ethiopia: Institutional-based prospective cohort study. PLoS One [Internet]. 2024;19(6.0):1–22. Available from: 10.1371/journal.pone.0302665

39. Getaneh FB, Belete AG, Ayres A, Ayalew T, Muche A, Derseh L. A generalized Poisson regression analysis of determinants of early neonatal mortality in Ethiopia using 2019 Ethiopian mini demographic health survey. Sci Rep [Internet]. 2024; 14(1):1–9. Available from: 10.1038/s41598-024-53332-5

40. Limaso A. A, Dangisso M. H, and Hibstu D. T, “Neonatal survival and determinants of mortality in Aroresa district, southern Ethiopia: a prospective cohort study,” BMC Pediatrics, vol. 20, no. 1, p. 33, 2020

41. Golubnitschaja O, Yeghiazaryan K, Cebioglu M, Morelli M, and Herrera-Marschitz M. Birth asphyxia as the major complication in newborns: moving towards improved individual outcomes by prediction, targeted prevention and tailored medical care. EPMA Journal. 2011; 2(2):197–210.

42. Ajaari J. Impact of place of delivery on neonatal mortality in rural Tanzania. Value in Health 2013; 16:A209–10.

43. Ronsmans C, Scott S, Qomariyah SN, et al. Professional assistance during birth and maternal mortality in two Indonesian districts. Bull World Health Organ 2009; 87:416– 23.

44. Mengesha, H. G., Lerebo, W. T., Kidanemariam, A., Gebrezgiabher, G. & Berhane, Y. Pre-term and post-term births: Predictors and implications on neonatal mortality in Northern Ethiopia. BMC Nurs. 15(1), 1–11 (2016).

45. Villar, J. et al. International standards for newborn weight, length, and head circumference by gestational age and sex: The Newborn Cross-Sectional Study of the INTERGROWTH-21st Project. Lancet 384(9946), 857–868 (2014).

46. Wastnedge E, Waters D, Murray SR, McGowan B, Chipeta E, Nyondo-Mipando AL, Gadama L, Gadama G, Masamba M, Malata M, Taulo F, Dube Q, Kawaza K, Khomani PM, Whyte S, Crampin M, Freyne B, Norman JE, Reynolds RM; DIPLOMATIC Collaboration. Interventions to reduce preterm birth and stillbirth, and improve outcomes for babies born preterm in low- and middle-income countries: A systematic review. J Glob Health. 2021 Dec 30; 11: 04050. doi: 10.7189/jogh.11.04050. PMID: 35003711; PMCID: PMC8709903.

47. Althabe F., et al., Cesarean section rates and maternal and neonatal mortality in low-, medium-, and high-income countries: an ecological study. Birth, 2006. 33(4): p. 270–277. 10.1111/j. 1523-536X.2006.00118.x PMID: 17150064

48. Gregory K.D., et al., Cesarean versus vaginal delivery: whose risks? Whose benefits? American journal of perinatology, 2012. 29(01): p. 07–18. 10.1055/s-0031-1285829 PMID: 21833896

49. Suparmi S., Chiera B., and Pradono J., Low birth weights and risk of neonatal mortality in Indonesia. Health Science Journal of Indonesia, 2016. 7(2): p. 113–117.

50. Migoto M.T., et al., Early neonatal mortality and risk factors: a case-control study in Paraná State. Revista Brasileira de Enfermagem, 2018. 71: p. 2527–2534

51. Soon B.T., The global action report on preterm birth. Geneva: World Health Organization, 2012: p. 2.

52. Conde-Agudelo A D.R.J. and Belizan J., Kangaroo mother care to reduce morbidity and mortality in low birthweight infants (Review). The Cochrane Collaboration. 2005, Wiley.

